# Urine pH and Kidney Outcomes in Biopsy–Proven Kidney Disease: Association With Medullary Cast Formation

**DOI:** 10.64898/2026.03.26.26349355

**Authors:** Kenji Tsuji, Naruhiko Uchida, Hiroyuki Nakanoh, Kazuhiko Fukushima, Haruhito A. Uchida, Shinji Kitamura, Jun Wada

## Abstract

**Background:** Lower urine pH has been associated with reduced kidney function and an increased risk of kidney disease; however, its prognostic and pathological significance in biopsy–proven kidney disease remains unclear. A recent study demonstrated that medullary cast formation is independently associated with adverse renal outcomes beyond established predictors such as interstitial fibrosis and tubular atrophy (IFTA), yet its clinical determinants are not fully elucidated. Urine pH reflects the intratubular acid–base microenvironment and may contribute to tubular obstruction through cast formation. In this study, we examined kidney outcomes in patients undergoing native kidney biopsy, and the associations of urine pH with medullary cast formation.

**Methods:** Among 1,167 adults who underwent native kidney biopsy between 2011 and 2024, 503 patients with evaluable medullary tissue were included in this retrospective observational cohort study. Urine pH was analyzed in relation to clinical and histological variables and kidney outcomes. The primary outcome was a ≥40% decline in estimated glomerular filtration rate (eGFR) or initiation of renal replacement therapy.

**Results:** The mean baseline eGFR was 54.3±17.7 mL/min/1.73 m², the mean urine pH was 6.15±0.84, and the median urinary protein excretion was 1.1 g/gCr. During a median follow-up of 2.11 years, 113 patients (22.5%) reached the kidney outcome. Kaplan–Meier analysis showed that lower urine pH was associated with a higher risk of kidney outcomes. In Cox proportional hazards models adjusted for proteinuria, baseline eGFR, and IFTA score, urine pH remained independently associated with kidney outcomes (hazard ratio, 0.69; 95% confidence interval, 0.51–0.91). Inclusion of urine pH improved prognostic discrimination beyond established risk factors (Harrell’s C-index, 0.642 to 0.654). Lower urine pH was also associated with greater medullary cast formation.

**Conclusion:** In patients undergoing native kidney biopsy, lower urine pH was independently associated with adverse kidney outcomes and greater medullary cast formation.

## Introduction

Chronic kidney disease (CKD) is a major global health burden, and identification of modifiable prognostic factors remains a critical issue in nephrology ^1^. Renal biopsy provides valuable histopathological information that contributes not only to diagnosis but also to prognostic stratification ^2^. While cortical lesions have been extensively studied as predictors of renal outcome, the prognostic significance of medullary pathological changes has received comparatively less attention. The renal medulla is particularly vulnerable to hypoxia and tubular obstruction ^3,4^, suggesting that medullary lesions may have important prognostic implications.

Among tubulointerstitial lesions, tubular cast formation, particularly within the renal medulla, has been reported as a potential indicator of adverse renal prognosis^5^. Cast formation is thought to reflect intratubular precipitation of proteins, cellular debris, or crystals, leading to tubular obstruction, increased intratubular pressure, and subsequent tubular injury ^5–8^. Recently, we reported that medullary cast formation is independently associated with kidney outcomes and provides prognostic value beyond established clinical and histological risk factors ^5^. These findings underscore the importance of the medullary microenvironment and tubular factors in determining long-term renal outcomes.

Urine pH is a readily available and repeatedly measurable clinical parameter that reflects renal acid–base handling and the intrarenal tubular microenvironment ^9^. Given its potential influence on protein aggregation and solubility, urine pH may play a role in modulating medullary cast formation. Experimental and clinical evidence suggests that acidic urine promotes protein aggregation and cast formation, whereas alkaline conditions may increase solubility and reduce intratubular obstruction ^6,10^. In addition, acidic tubular conditions have been linked to tubular injury and profibrotic responses ^11^. Epidemiological studies suggest that lower urine pH is associated with kidney function decline across diverse populations, including CKD ^12–16^. However, the pathological mechanisms underlying this association remain incompletely understood, particularly with respect to medullary cast formation in biopsy-proven kidney disease.

We therefore hypothesized that urine pH influences renal prognosis through modulation of medullary cast formation, thereby contributing to tubular obstruction and subsequent renal function decline. To test this hypothesis, we conducted a cohort study of patients undergoing native kidney biopsy, examining the association between urine pH, medullary cast formation, and long-term renal outcomes. Elucidating this relationship may provide novel insights into the pathophysiology of CKD progression and identify urine pH as a potential, modifiable prognostic marker in patients with biopsy-proven kidney disease.

## Materials and Methods

### Study Design and participants

We retrospectively reviewed 1,136 adult patients who underwent a native kidney biopsy in the Okayama University Hospital (Okayama, Japan) between January 2011 and March 2024. The exclusion criteria were eGFR ≤15 mL/min/1.73 m², dialysis at the time of biopsy, age <18 years, or insufficient medullary tissue. Consequently, 503 patients were enrolled in the study (Figure 1). The study protocol was approved by the ethics committee of Okayama University Hospital Institutional Review Board (accredited ISO9001/2000), Okayama, Japan (approval number: 1908-022 and 2407-038), and is in accordance with the principles of the Declaration of Helsinki. The requirement for written informed consent was waived due to the retrospective nature of the study. Instead, information about the study was publicly disclosed on the institutional website, and patients were provided the opportunity to opt out. This study was conducted in an expanded cohort that included additional cases beyond our previous analyses.

**Figure 1.**
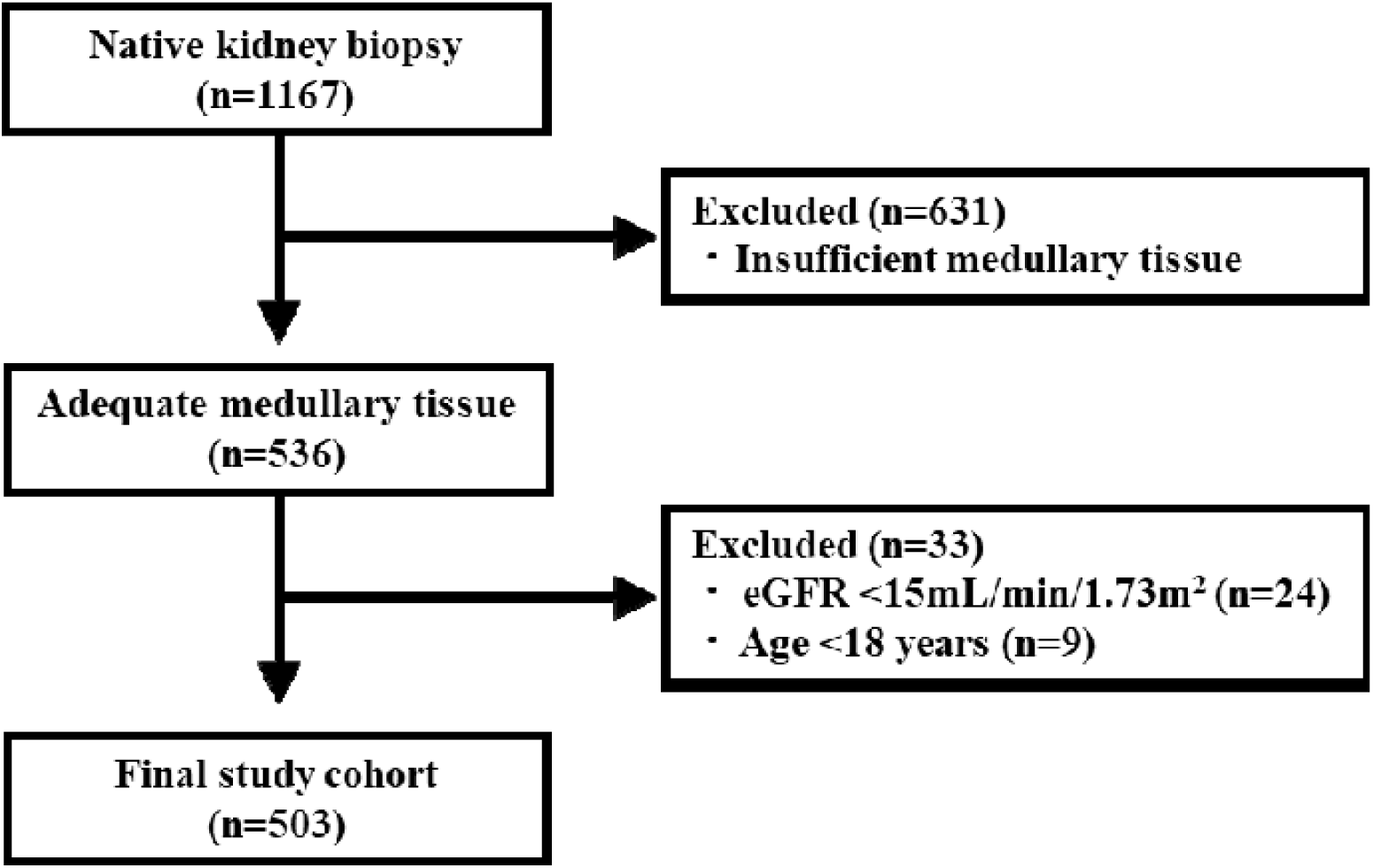
Flowchart of study participants. eGFR, estimated glomerular filtration rate.

### Histological parameters

Obtained kidney tissues were processed for light microscopy, immunofluorescent staining and electron microscopy. For light microscopy, specimens were stained with hematoxylin and eosin, periodic acid-Schiff, periodic acid-methenamine silver and Masson’s trichrome, according to routine methods. All biopsies were adjudicated to determine histologic scores and diagnosis by at least two experienced nephrologists, through conference discussions based on patients’ medical histories, physical information, clinical examinations and pathological findings. The IFTA score were graded on a scale of 0, 1, 2 and 3 (≤10%, 11-25%, 26-50%, >50%, respectively) according to previous reports and the Banff classification of kidney allograft pathology ^17^. Medullary fibrosis and cast formation and inflammation were scored as 0 (<10%), 1 (10–24%), or 2 (≥25%), and inflammatory cell infiltration was scored as 0 (<5%), 1 (5–14%), or 2 (≥15%) (Supplemental Figure 1), as previously reported ^5^.

### Clinical Parameters

The following clinicopathologic characteristics were collected at the time of kidney biopsies: age, sex, body mass index (BMI), hemoglobin A1c (HbA1c), the use of angiotensin-converting-enzyme inhibitor (ACE-I) and/or angiotensin II receptor blocker (ARB), blood pressure, history of smoking, serum creatinine (s-Cr) and eGFR levels, urine pH, urinary protein levels and hematuria (>5 erythrocytes per high power field). eGFR was evaluated using the equation developed by the Japanese Society of Nephrology ^18^. Hypertension was defined as blood pressure (BP) ≥140/90 mmHg at the time of the kidney biopsy or use of antihypertensive drugs. HbA1c data are presented as National Glycohemoglobin Standardization Program values according to the recommendations of the Japanese Diabetes Society and the International Federation of Clinical Chemistry ^19^.

### Exposure and Outcomes

The primary exposure was urine pH. Urine pH was categorized into tertiles based on its distribution (pH<6.0, 6-<7.0, 7.0<). Established prognostic factors, including IFTA scores, were included as covariates ^2^. The primary endpoint was defined as an eGFR decline of ≥40% or the commencement of renal replacement therapy (dialysis or kidney transplantation) ^20^. eGFR levels were obtained from electronic medical records during follow-up.

### Statistical analysis

Data were summarized as percentages for categorical variables and means ±□standard deviation (SD) or median (interquartile range) for continuous variables. Skewed variable (proteinuria) underwent a natural logarithmic transformation before the analysis. Categorical variables were analyzed using a chi-squared test, whereas continuous variables were compared using Student’s t-test or a Mann–Whitney U test, as appropriate. *p* for trend was calculated by the Cochran–Armitage trend test or the Jonckheere–Terpstra test for the distribution of each clinical parameter, stratified by IFTA score. Kidney survival was estimated using the Kaplan–Meier method and compared by the log-rank test. Patients were censored at the last follow-up or October 31, 2024. Cox proportional hazards models were used to estimate hazard ratios (HRs) and 95% confidence intervals (CIs), with adjustment for covariates selected based on biological plausibility and prior reports. Discriminative ability was assessed by Harrell’s concordance index (C-index) ^21^. Correlations between urine pH and clinical variables were evaluated using Spearman’s correlation analysis. To evaluate factors associated with urine pH, we performed ordinal logistic regression analyses using categorized urine pH levels, and multiple linear regression analyses treating urine pH as a continuous variable. Candidate variables included age, sex, eGFR, diabetes, hypertension, BMI, urinary protein levels, ACE-I or ARB use, and hematuria. K Statistical analyses were performed using JMP version 17.0.0 (SAS Institute, Inc., Cary, NC). All tests were two-sided, and p < 0.05 was considered statistically significant.

## Results

### Patient characteristics at the time of kidney biopsy

Among 1,167 patients who underwent a kidney biopsy, 503 patients met the selection criteria and were enrolled in the analysis (Figure 1). Clinical and histopathological variables at the time of kidney biopsy, stratified by urine pH and IFTA score, as well as primary clinicopathologic diagnoses were summarized (Table 1, Supplemental Table 1, Supplemental Table 2). The mean age was 54.3±17.7 years; 222 patients (44%) were male, 126 patients (25%) had a history of diabetes, 235 patients (46.7%) had a history of hypertension, the mean eGFR was 60.7 ± 26.2 ml/min per 1.73 m^2^, and urine pH was 6.15 ± 0.84, and median proteinuria was 1.1 g/gCr (interquartile range (IQR), 0.37–3.1). Patients with lower urine pH had higher serum creatinine levels, higher IFTA scores, more severe medullary fibrosis and cast formation, and a higher prevalence of hypertension and diabetes. They also had lower eGFR and BMI.

**Table 1.** Baseline clinical and histopathological characteristics according to urine pH score.

|  |  | All patients<br>(n=503) | Urine pH score |  |  | p for<br>trend |
| --- | --- | --- | --- | --- | --- | --- |
|  |  |  | 0 (n=192) | 1 (n=186) | 2 (n=125) |  |
| Men |  | 222 (44.0%) | 95 (49.5%) | 84 (45.2%) | 44 (35.2%) | 0.016 |
| Age, years |  | 54.3±17.7 | 56.9±16.2 | 54.2±18.8 | 50.2±17.9 | 0.003 |
| BMI, kg/m<sup>2</sup> |  | 22.7±4.0 | 23.1±4.3 | 22.9±3.7 | 21.8±3.7 | 0.009 |
| sBP, mmHg |  | 127.3±18.6 | 128.7±19.2 | 128.7±17.6 | 123.2±18.6 | 0.015 |
| dBP, mmHg |  | 79.4±12.5 | 80.1±13.0 | 78.9±11.7 | 78.8±12.8 | 0.41 |
| sCr, mg/dL |  | 1.06±0.49 | 1.21±0.56 | 1.07±0.46 | 0.83±0.32 | <0.001 |
| eGFR, ml/min/1.73 m<sup>2</sup> |  | 60.7±26.2 | 52.9±22.8 | 60.2±26.8 | 73.9±25.5 | <0.001 |
| HbA1c, % |  | 5.7 (5.4-6.1) | 5.8 (5.5-6.2) | 5.7 (5.4-6.2) | 5.6 ( 5.3-5.9) | 0.001 |
| U-P, g/gCr |  | 1.1<br>(0.37-3.1) | 0.98<br>(0.37-2.1) | 1.4<br>(0.44-4.0) | 1.0<br>(0.31-3.4) | 0.21 |
| Hematuria, % |  | 296 (58.8%) | 113 (58.9%) | 113 (60.8%) | 70 (56.0%) | 0.71 |
| Urine pH |  | 6.15±0.84 | 5.40±0.20 | 6.20±0.25 | 7.36±0.44 | <0.001 |
| Urine cast |  | 233 (46.3%) | 106 (55.2%) | 90 (48.4%) | 38 (30.4%) | <0.001 |
| Hypertension |  | 235 (46.7%) | 102 (53.1%) | 89 (47.8%) | 44 (35.2%) | 0.003 |
| DM |  | 126 (25.0%) | 54 (28.1%) | 51 (27.4%) | 21 (16.8%) | 0.040 |
| ACE-I or ARB |  | 175 (34.8%) | 87 (45.3%) | 62 (33.3%) | 26 (3.7%) | <0.001 |
| Ca blocker |  | 176 (35.0%) | 74 (38.5%) | 66 (35.5%) | 36 (28.8%) | 0.087 |
| Statins |  | 121 (24.1%) | 47 (24.5%) | 44 (23.7%) | 30 (24.0%) | 0.87 |
| IFTA | 0 | 85 (16.9%) | 20 (10.4%) | 27 (14.5%) | 38 (30.4%) | <0.001 |
|  | 1 | 276 (54.9%) | 97 (50.5%) | 111 (59.7%) | 69 (55.2%) |  |
|  | 2 | 132 (26.2%) | 73 (38.0%) | 42 (22.6%) | 17 (13.6%) |  |
|  | 3 | 9 (1.8%) | 2 (1.0%) | 6 (3.2%) | 1 (0.8%) |  |
| Medullary<br>fibrosis | 0 | 138 (27.4%) | 43 (22.4%) | 53 (28.5%) | 42 (33.6%) | 0.001 |
|  | 1 | 247 (49.1%) | 94 (49.0%) | 86 (46.2%) | 68 (54.4%) |  |
|  | 2 | 117 (23.3%) | 55 (28.6%) | 47 (25.3%) | 15 (12.0%) |  |
| Medullary<br>inflammatory cell<br>infiltration | 0 | 366 (72.8%) | 131 (68.2%) | 137 (73.7%) | 99 (79.2%) | 0.049 |
|  | 1 | 111 (22.1%) | 54 (28.1%) | 36 (19.4%) | 21 (16.8%) |  |
|  | 2 | 25 (5.0%) | 7 (3.6%) | 13 (7.0%) | 5 (4.0%) |  |
| Medullary cast<br>formation | 0 | 258 (51.3%) | 80 (41.7%) | 95 (51.1%) | 84 (67.2%) | <0.001 |
|  | 1 | 178 (35.4%) | 80 (41.7%) | 66 (35.5%) | 32 (25.6%) |  |
|  | 2 | 66 (13.1%) | 32 (16.7%) | 25 (13.4%) | 9 (7.2%) |  |

Values reported as n (%) for categorical variables and mean ± standard deviation or median (interquartile range) for continuous variables. BMI, body mass index; sBP, systolic blood pressure; dBP, diastolic blood pressure; sCr, serum creatinine; eGFR, estimated glomerular filtration rate; HbA1c, hemoglobin A1c; U-P, urinary protein excretion; DM, diabetes mellitus; ACE-I, angiotensin-converting enzyme inhibitor; ARB, angiotensin II type 1 receptor blocker; IFTA, interstitial fibrosis and tubular atrophy.

### Prognostic significance of histopathologic lesions

During the median follow-up period of 2.11 years, 113 patients (22.5%) experienced the composite kidney outcome. Kaplan–Meier analyses showed that higher IFTA scores were associated with an increased risk of kidney outcomes. Similarly, compared with the intermediate urine pH group, kidney survival was lowest in the low urine pH group and highest in the high urine pH group (Figure 2). In Cox proportional hazards models adjusted for age, gender, eGFR and proteinuria, both urine pH categories and IFTA scores were independently associated with kidney outcomes (Table 2, Figure 3). This association remained significant after further adjustment for IFTA score (hazard ratio [HR], 0.71; 95% confidence interval [CI], 0.53–0.95). Similar trends were observed when urine pH was analyzed as a continuous variable (HR, 0.69; 95% CI, 0.51–0.91) (Table 2).

**Figure 2.**
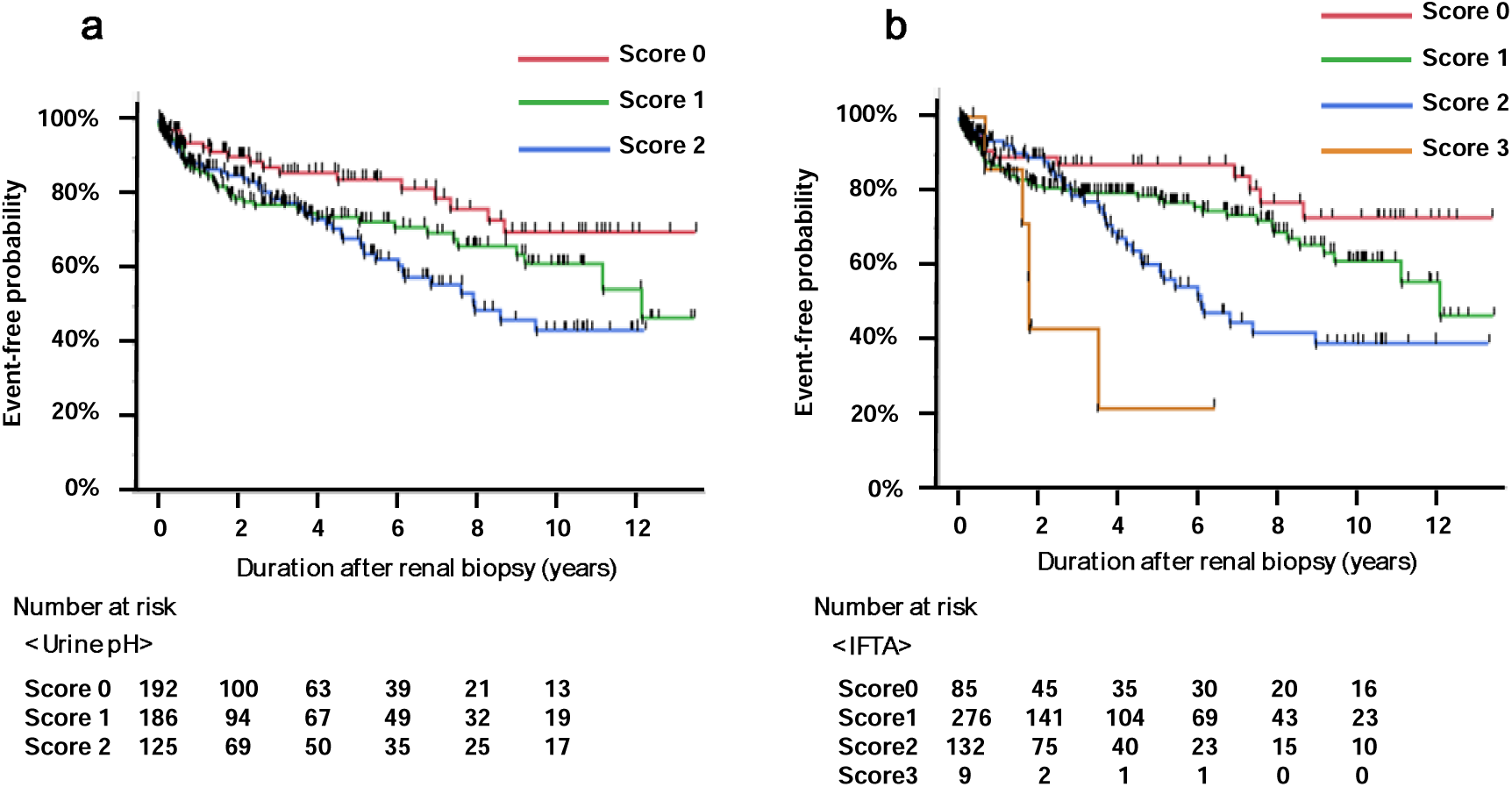
Renal survival rate stratified by urine pH and IFTA score. (a) Renal survival rate stratified by urine pH score. Log rank test: *p* = 0.019. (b) Renal survival rate stratified IFTA score. Log rank test: *p* <0.001. IFTA, interstitial fibrosis and tubular atrophy.

**Figure 3.**
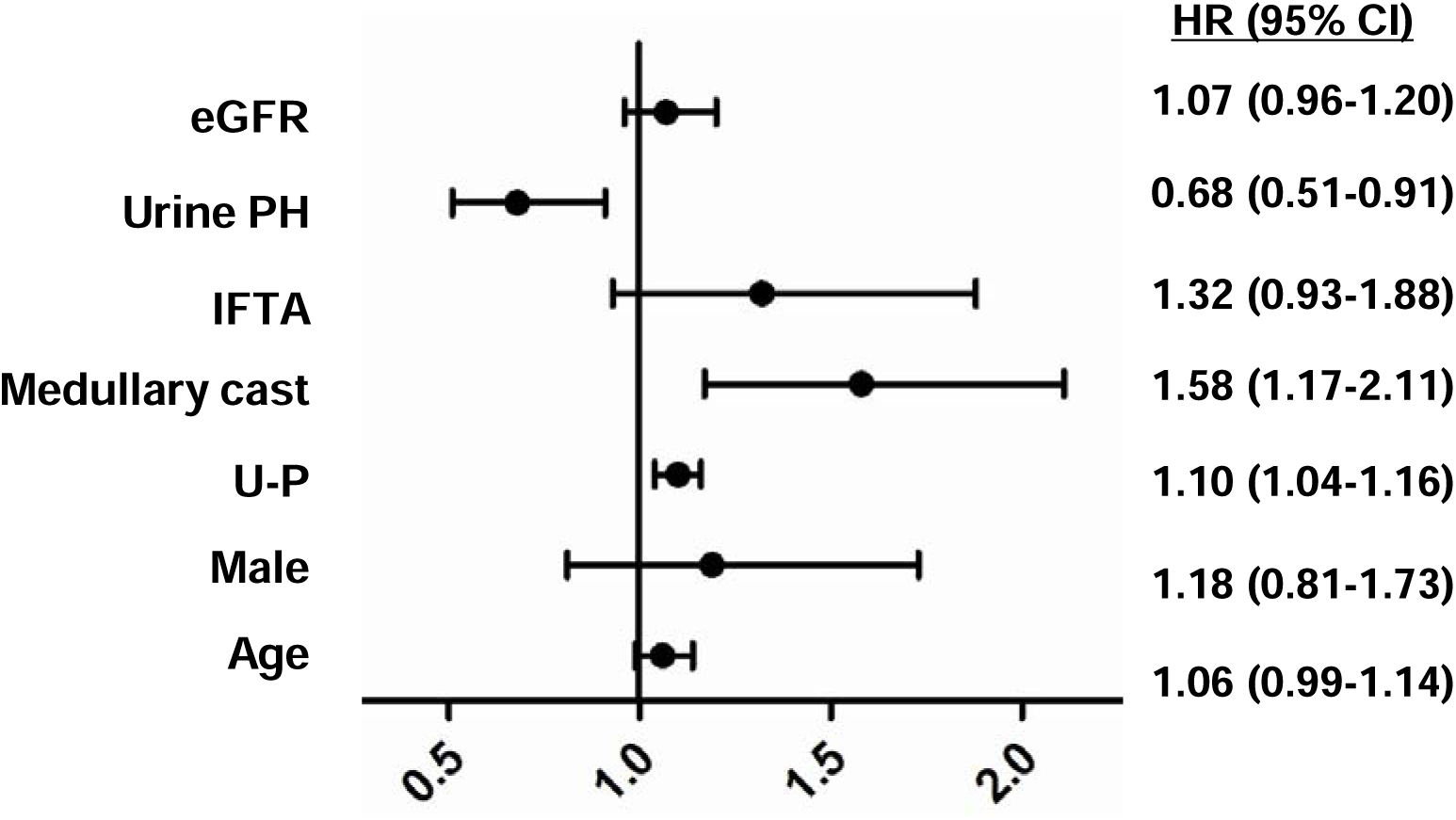
Multivariable Cox proportional hazards models for kidney outcomes. Forest plot showing hazard ratios and 95% confidence intervals for the associations of urine pH, eGFR (per 10 mL/min/1.73 m² decrease), interstitial fibrosis and tubular atrophy (IFTA) score, medullary fibrosis score, urine protein (U-P), age (per 5-year increase), and sex with kidney outcomes.

**Table 2.** Multivariable Cox proportional hazards models for kidney outcomes.

|  | Model 0<br>HR (95%CI) | Model 1<br>HR (95%CI) | Model 2<br>HR (95%CI) | Model 3-1<br>HR (95%CI) | Model 3-2<br>HR (95%CI) | Model 3-3<br>HR (95%CI) |
| --- | --- | --- | --- | --- | --- | --- |
| Urine pH<br>score<br>(per score) | 0.70<br>(0.55-0.90) | 0.76<br>(0.59-0.98) | 0.68<br>(0.51-0.91) | 0.71<br>(0.53-0.95) | 0.68<br>(0.50-0.90) | 0.68<br>(0.51-0.91) |
| Urine pH<br>(continuous) | 0.68<br>(0.53-0.87) | 0.73<br>(0.56-0.94) | 0.66<br>(0.49-0.88) | 0.69<br>(0.51-0.91) | 0.66<br>(0.49-0.87) | 0.67<br>(0.50-0.89) |

Model 0: No adjustment factors. Model 1: Sex, Age. Model 2: Model 1 + Urinary protein, eGFR. Model 3-1: Model 2 + IFTA. Model 3-2: Model 2 + medullary fibrosis. Model 3-3: Model 2 + medullary cast formation. HR, hazard ratio; 95%CI, 95% confidence interval; IFTA, interstitial fibrosis and tubular atrophy.

To assess whether medullary pathological findings improved prognostic discrimination beyond conventional risk factors, the C-index was calculated. A model including age, sex, eGFR, and proteinuria yielded a C-index of 0.632 (standard error [SE], 0.030). Addition of cortical IFTA increased the C-index to 0.642 (SE, 0.028), and further inclusion of urine PH increased it to 0.654 (SE, 0.028), indicating incremental improvement in prognostic performance.

In a subgroup analysis of patients with IgA nephropathy (n = 158), 28 kidney outcome events occurred during a median follow-up of 3.1 years. Kaplan–Meier analysis showed a similar trend to that observed in the overall cohort, although the differences were not statistically significant (Supplemental Figure 2). Multivariable Cox analysis was not performed due to the limited number of events.

### Predictors of urine pH

To explore predictors of lower urine pH, patients were stratified according to the underlying kidney disease. Mean urine pH varied across disease categories, with relatively higher values observed in minimal change disease and membranous nephropathy, and lower values in diabetic nephropathy and nephrosclerosis, indicating heterogeneity across disease etiologies (Supplemental Figure 3). (Supplemental Figure 3). Next, factors associated with urine pH were evaluated using Spearman’s correlation and ordinal logistic regression analyses (Table 3 and Supplemental Table 4). Urine pH was positively correlated with eGFR (Spearman’s ρ = 0.30, p < 0.001) and inversely correlated with medullary cast score (ρ = −0.19, p < 0.001) and IFTA score (ρ = −0.25, p < 0.001). Correlations between urine pH and clinical variables are shown in Supplemental Figure 4. In ordinal logistic regression analysis, lower eGFR, higher urine protein, and IFTA were significantly associated with lower urine pH. Male sex was also associated with urine pH, although the effect size was modest. Age was not significantly associated.

**Table 3.**
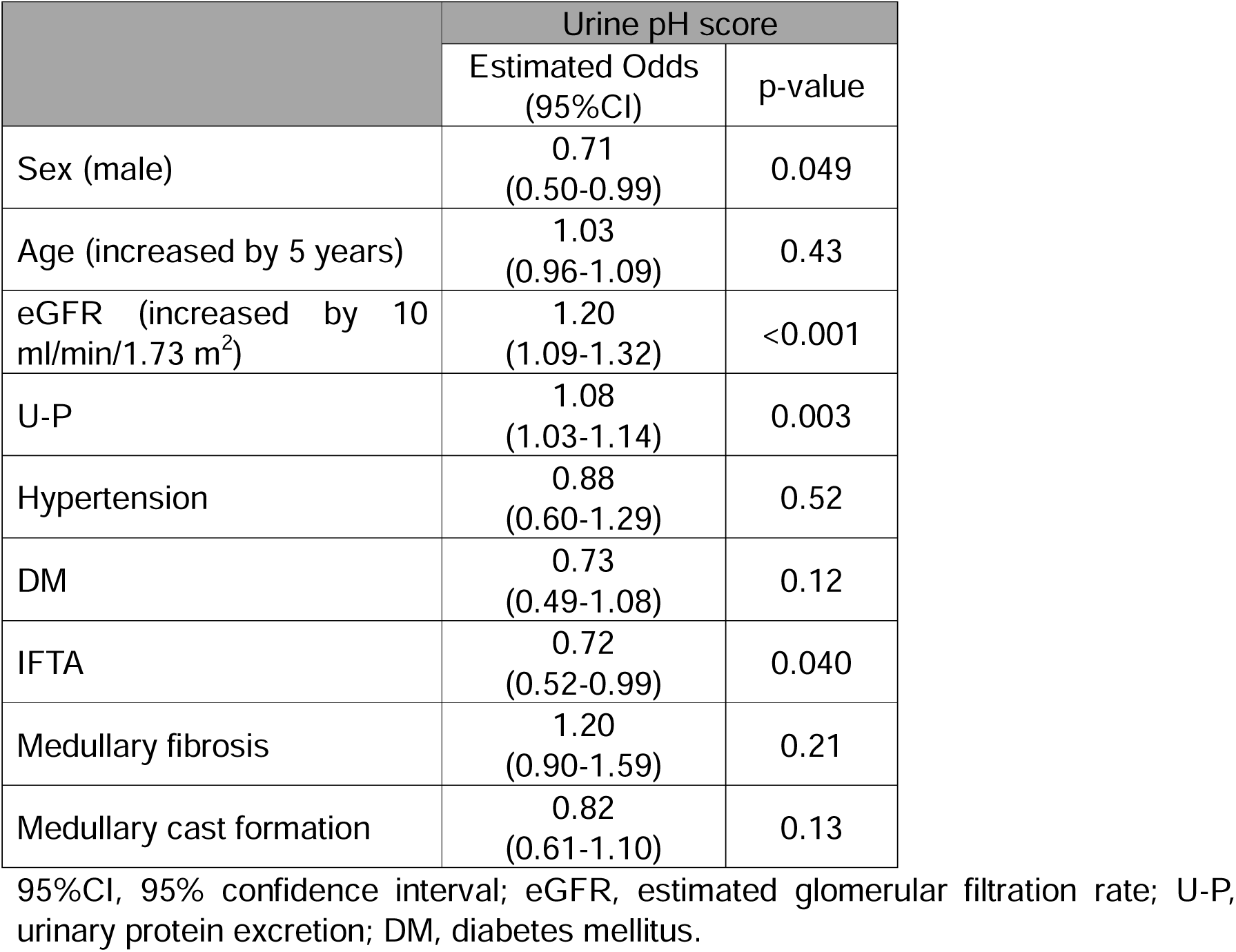
Ordinal logistic regression analysis by urine PH score.

|  | Urine pH score |  |
| --- | --- | --- |
|  | Estimated Odds<br>(95%CI) | p-value |
| Sex (male) | 0.71<br>(0.50-0.99) | 0.049 |
| Age (increased by 5 years) | 1.03<br>(0.96-1.09) | 0.43 |
| eGFR (increased by 10<br>ml/min/1.73 m<sup>2</sup>) | 1.20<br>(1.09-1.32) | <0.001 |
| U-P | 1.08<br>(1.03-1.14) | 0.003 |
| Hypertension | 0.88<br>(0.60-1.29) | 0.52 |
| DM | 0.73<br>(0.49-1.08) | 0.12 |
| IFTA | 0.72<br>(0.52-0.99) | 0.040 |
| Medullary fibrosis | 1.20<br>(0.90-1.59) | 0.21 |
| Medullary cast formation | 0.82<br>(0.61-1.10) | 0.13 |

95%CI, 95% confidence interval; eGFR, estimated glomerular filtration rate; U-P, urinary protein excretion; DM, diabetes mellitus.

## Discussion

In this biopsy-proven CKD cohort, lower urine pH was independently associated with adverse kidney outcomes, even after adjustment for established prognostic factors, including baseline eGFR, proteinuria, and IFTA score. In addition, urine pH was associated with medullary cast formation, suggesting a potential link between urinary acidification and tubular pathology (Figure 4). These findings provide clinicopathological insight into the role of the intrarenal acid–base microenvironment in CKD progression.

**Figure 4.**
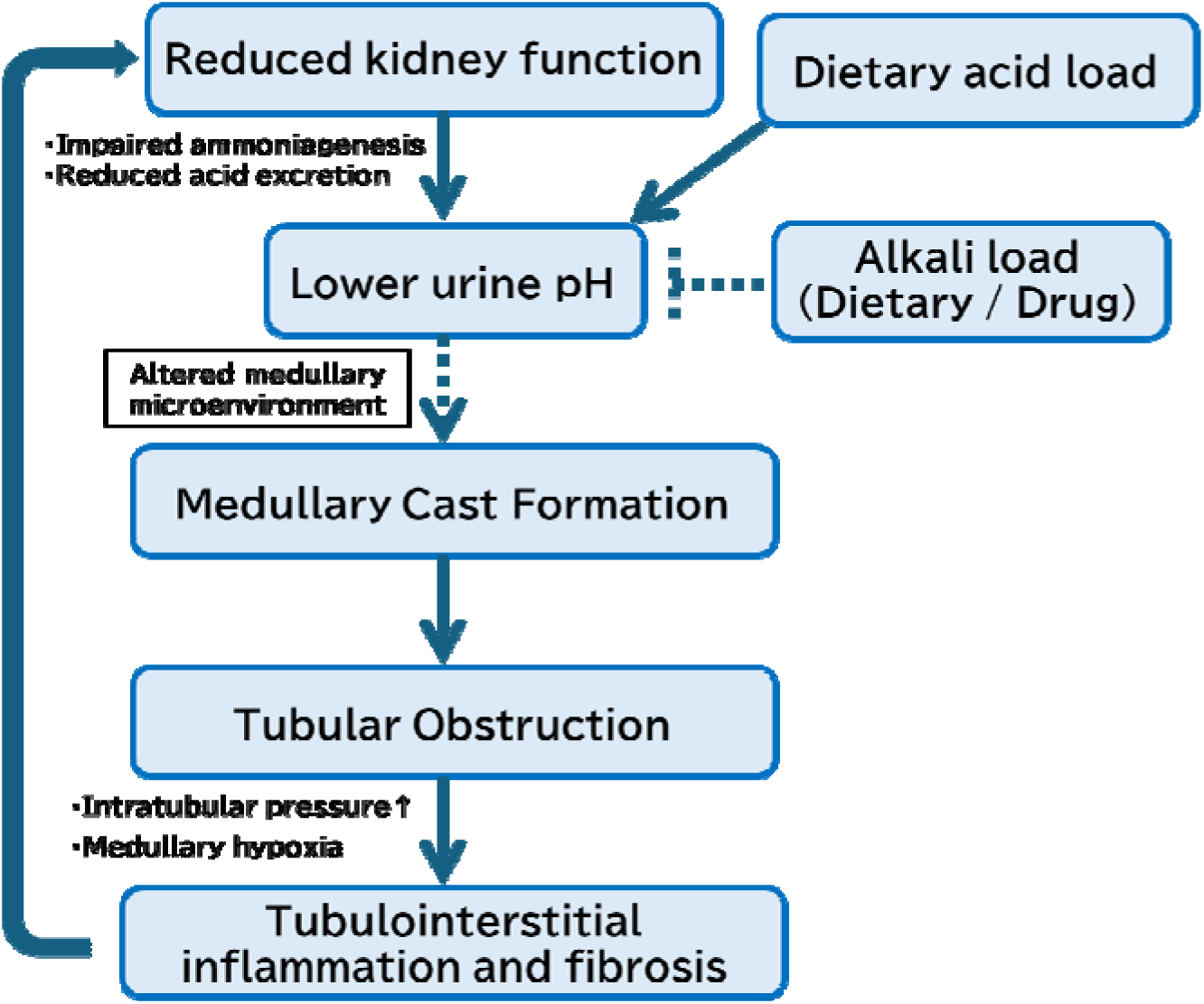
Conceptual model linking urinary acidification, medullary cast formation, and kidney function decline. Proposed framework illustrating how reduced kidney function and increased dietary acid load may contribute to urinary acidification, which in turn may promote medullary cast formation, tubular obstruction, and tubulointerstitial injury. These processes may further accelerate kidney function decline, forming a bidirectional pathogenic loop. Dietary or pharmacologic alkali supplementation is shown as a potentially modifiable factor that may mitigate urinary acidification. This model is based on the findings of the present study.

Urine pH reflects renal acid excretion and intratubular hydrogen ion handling^9^. Previous studies have reported that lower urine pH is associated with adverse kidney outcomes, including CKD progression, across diverse populations. ^12–16^. Our findings extend these observations to a biopsy-based cohort, enabling direct evaluation of histopathological correlates. Importantly, urine pH retained prognostic significance even after adjustment for IFTA, suggesting that it captures information beyond static structural injury and may reflect ongoing tubular stress or maladaptive responses. Experimental and clinical studies provide mechanistic support for a role of urinary acid–base status in tubular injury ^6,10,11^. Dietary alkali intake and alkali supplementation have been associated with slower CKD progression and limitation of oxidative stress, potentially through improvement of the intratubular environment^22–24^, supporting the concept that modulation of intrarenal acid–base balance has therapeutic relevance.

A key observation of this study is the association between lower urine pH and medullary cast formation. Acidic tubular conditions may reduce the solubility of filtered proteins and promote aggregation within the medullary environment, potentially contributing to tubular obstruction. Indeed, experimental study supports this concept ^25^, demonstrating that alkalinization reduces tubular cast formation and tubulointerstitial injury. On the other hand, the association between urine pH and medullary cast formation was attenuated after multivariable adjustment. This finding suggests that urine pH may not act as an independent determinant of cast formation, but rather reflects one aspect of a complex pathophysiological process involving reduced kidney function, tubular injury, and altered intrarenal microenvironment. Reduced kidney function may promote urinary acidification, while cast formation may further contribute to tubular injury and functional decline. Thus, urine pH may function as an integrative marker linking kidney function and tubular pathology. From a clinical perspective, urine pH is a simple, readily available, repeatedly measurable, and potentially modifiable clinical parameter. Alkali supplementation and dietary interventions have been reported to slow CKD progression ^22,26^. However, our findings suggest that targeting urinary acidification alone may be insufficient to fully prevent medullary cast formation, and that a multifaceted approach addressing additional determinants of tubular injury may be required (Figure 4).

This study has several limitations. First, this was a retrospective single-center study, which may limit generalizability and introduce selection bias. Second, urine pH was measured at a single time point and may have been influenced by dietary factors and medications, potentially leading to misclassification of exposure. Third, urinary ammonium excretion was not assessed, precluding a more comprehensive evaluation of renal acid–base handling. Fourth, biopsy sampling may not fully capture medullary pathology because of anatomical heterogeneity and limited tissue availability. Finally, as an observational study, causal relationships cannot be established.

In conclusion, lower urine pH was independently associated with adverse kidney outcomes and with medullary cast formation in a biopsy–proven cohort. These findings suggest that urinary acidification may contribute to tubular injury and CKD progression. Urine pH is a simple, readily available, and potentially modifiable clinical parameter that may provide insight into intrarenal pathophysiology and identify a potential therapeutic target.

## Data Availability

All data produced in the present study are available upon reasonable request to the authors

## Acknowledgements

The authors used ChatGPT (OpenAI, San Francisco, CA, USA) for English language editing and manuscript refinement. The authors take full responsibility for the content of this manuscript.

## Author contributions

K. Tsuji designed the study and drafted the paper; K. Tsuji, N. Uchida, H. Nakanoh, and K. Fukushima prepared and organized study datasets; K. Tsuji, N. Uchida, H. Nakanoh analyzed the data; K. Tsuji and N. Uchida made the figures; HA. Uchida, S. Kitamura and J. Wada reviewed statistical methods and analysis; all authors revised and approved the final version of the manuscript.

## Data Sharing Statement

The data that support the findings of this study are available from the corresponding author upon reasonable request.

## Supplementary Material

**Supplemental Table S1.** Clinical and histopathological findings for all patients and for patients stratified by IFTA score.

**Supplemental Table S2.** Clinicopathologic diagnoses of the study population.

**Supplemental Table S3.** Cox proportional hazards analysis of urine pH score.

**Supplemental Table S4.** Spearman’s correlation coefficients among clinical and pathological variables.

**Supplemental Figure S1.** Medullary pathological findings and scoring system.

**Supplemental Figure S2.** Renal survival rate stratified by urine pH and IFTA score in IgA nephropathy.

**Supplemental Figure S3.** Urine pH according to underlying kidney disease.

**Supplemental Figure S4.** Correlation between urine pH and clinical variables.

