## Supplemental Materials for "Urine pH and Kidney Outcomes in Biopsy–Proven Kidney Disease: Association With Medullary Cast Formation"

### Supplemental Material

#### Table of contents


**1. Supplemental Table**

**Supplemental Table 1.** Clinical and histopathological findings for all patients and for patients stratified by IFTA score

**Supplemental Table 2.** Clinicopathologic diagnoses of the study population

**Supplemental Table 3.** Cox proportional hazards analysis of urine pH score

**Supplemental Table 4.** Spearman's correlation coefficients among clinical and pathological variables

**2. Supplemental Figure**

**Supplemental Figure 1.** Medullary pathological findings and scoring system.

**Supplemental Figure 2.** Renal survival rate stratified by urine pH and IFTA score in IgA nephropathy.

**Supplemental Figure 3.** Urine pH according to underlying kidney disease.

**Supplemental Figure 4.** Correlation between urine pH and clinical variables.

**3. STROBE Statement**

**Supplemental Table 1.** Clinical and histopathological findings for all patients and for patients stratified by IFTA score

<table>
<tr><th rowspan="2" colspan="2"></th><th rowspan="2">All patients (n=503)</th><th colspan="4">IFTA score</th><th rowspan="2">p for trend</th></tr>
<tr><th>0 (n=85)</th><th>1 (n=277)</th><th>2 (n=132)</th><th>3 (n=9)</th></tr>
<tr><td colspan="2">Men</td><td>222 (44.0%)</td><td>37 (43.5%)</td><td>129 (46.6%)</td><td>52 (39.4%)</td><td>5 (55.6%)</td><td>0.52</td></tr>
<tr><td colspan="2">Age, years</td><td>54.3±17.7</td><td>41.6±18.0</td><td>54.9±17.0</td><td>60.8±14.9</td><td>55.6±18.0</td><td><0.001</td></tr>
<tr><td colspan="2">BMI, kg/m$^2$</td><td>22.7±4.0</td><td>22.9±4.2</td><td>22.7±3.7</td><td>22.4±4.5</td><td>23.5±3.9</td><td>0.58</td></tr>
<tr><td colspan="2">sBP, mmHg</td><td>127.3±18.6</td><td>120.4±15.8</td><td>127.0±18.9</td><td>131.9±18.3</td><td>135.7±17.9</td><td><0.001</td></tr>
<tr><td colspan="2">dBP, mmHg</td><td>79.4±12.5</td><td>77.5±10.5</td><td>79.5±12.1</td><td>80.3±14.1</td><td>75.9±15.6</td><td>0.28</td></tr>
<tr><td colspan="2">sCr, mg/dL</td><td>1.06±0.49</td><td>0.78±0.27</td><td>0.96±0.40</td><td>1.41±0.55</td><td>1.90±0.50</td><td><0.001</td></tr>
<tr><td colspan="2">eGFR, ml/min/1.73 m$^2$</td><td>60.7±26.2</td><td>84.6±26.5</td><td>63.9±22.2</td><td>41.2±17.8</td><td>28.9±7.66</td><td><0.001</td></tr>
<tr><td colspan="2">HbA1c, %</td><td>5.7 (5.4-6.1)</td><td>5.5 (5.3-5.8)</td><td>5.8 (5.5-6.2)</td><td>5.8 (5.4-6.1)</td><td>6.1 (5.1-6.9)</td><td>0.007</td></tr>
<tr><td colspan="2">U-P, g/gCr</td><td>1.10 (0.37-3.10)</td><td>0.70 (0.16-3.50)</td><td>1.00 (0.37-2.88)</td><td>1.33 (0.57-3.04)</td><td>4.99 (2.74-5.71)</td><td>0.004</td></tr>
<tr><td colspan="2">Hematuria, %</td><td>296 (58.8%)</td><td>44 (51.8%)</td><td>178 (64.3%)</td><td>67 (50.8%)</td><td>7 (77.8%)</td><td>0.66</td></tr>
<tr><td colspan="2">Urine pH</td><td>6.15±0.84</td><td>6.59±0.91</td><td>6.20±0.77</td><td>5.91±0.55</td><td>6.22±0.62</td><td><0.001</td></tr>
<tr><td colspan="2">Urine cast</td><td>233 (46.3%)</td><td>27 (31.8%)</td><td>133 (48.0%)</td><td>68 (51.5%)</td><td>6 (66.7%)</td><td>0.005</td></tr>
<tr><td colspan="2">Hypertension</td><td>235 (46.7%)</td><td>21 (24.7%)</td><td>124 (44.8%)</td><td>85 (64.4%)</td><td>5 (55.6%)</td><td><0.001</td></tr>
<tr><td colspan="2">DM</td><td>126 (25.0%)</td><td>16 (18.8%)</td><td>69 (24.9%)</td><td>36 (27.3%)</td><td>5 (55.6%)</td><td>0.069</td></tr>
<tr><td colspan="2">ACE-I or ARB</td><td>175 (34.8%)</td><td>11 (12.9%)</td><td>103 (37.2%)</td><td>58 (43.9%)</td><td>3 (33.3%)</td><td><0.001</td></tr>
<tr><td colspan="2">Ca blocker</td><td>176 (35.0%)</td><td>16 (18.8%)</td><td>87 (31.4%)</td><td>67 (50.8%)</td><td>6 (66.7%)</td><td><0.001</td></tr>
<tr><td colspan="2">Statins</td><td>121 (24.1%)</td><td>16 (18.8%)</td><td>71 (25.6%)</td><td>31 (23.5%)</td><td>3 (33.3%)</td><td>0.60</td></tr>
<tr><td rowspan="3">Medullary fibrosis</td><td>0</td><td>138 (27.4%)</td><td>63 (74.1%)</td><td>69 (24.9%)</td><td>6 (4.5%)</td><td>0 (0.0%)</td><td rowspan="3"><0.001</td></tr>
<tr><td>1</td><td>247 (49.1%)</td><td>20 (23.5%)</td><td>164 (59.2%)</td><td>63 (47.7%)</td><td>1 (11.1%)</td></tr>
<tr><td>2</td><td>117 (23.3%)</td><td>2 (2.4%)</td><td>44 (15.9%)</td><td>63 (47.7%)</td><td>8 (88.9%)</td></tr>
<tr><td rowspan="3">Medullary inflammatory cell infiltration</td><td>0</td><td>366 (72.8%)</td><td>81 (95.3%)</td><td>220 (79.4%)</td><td>64 (48.5%)</td><td>2 (22.2%)</td><td rowspan="3"><0.001</td></tr>
<tr><td>1</td><td>111 (22.1%)</td><td>4 (4.7%)</td><td>54 (19.5%)</td><td>51 (38.6%)</td><td>2 (22.2%)</td></tr>
<tr><td>2</td><td>25 (5.0%)</td><td>0 (0.0%)</td><td>3 (1.1%)</td><td>17 (12.9%)</td><td>5 (55.6%)</td></tr>
<tr><td rowspan="3">Medullary cast formation</td><td>0</td><td>258 (51.3%)</td><td>78 (91.8%)</td><td>153 (55.2%)</td><td>27 (20.5%)</td><td>1 (11.1%)</td><td rowspan="3"><0.001</td></tr>
<tr><td>1</td><td>178 (35.4%)</td><td>7 (8.2%)</td><td>98 (35.4%)</td><td>71 (53.8%)</td><td>2 (22.2%)</td></tr>
<tr><td>2</td><td>66 (13.1%)</td><td>0 (0.0%)</td><td>26 (9.4%)</td><td>34 (25.8%)</td><td>6 (66.7%)</td></tr>
</table>

Values reported as n (%) for categorical variables and mean ± standard deviation or median (interquartile range) for continuous variables. BMI, body mass index; sBP, systolic blood pressure; dBP, diastolic blood pressure; sCr, serum creatinine; eGFR, estimated glomerular filtration rate; HbA1c, hemoglobin A1c; U-P, urinary protein excretion; DM, diabetes mellitus; ACE-I, angiotensin–converting enzyme inhibitor; ARB, angiotensin II type 1 receptor blocker; IFTA, interstitial fibrosis and tubular atrophy.

**Supplemental Table 2. Clinicopathologic diagnoses of the study population**

| Clinicopathologic diagnosis | n=503 |
|---|---|
| IgA nephropathy | 158 |
| ANCA-associated vasculitis | 50 |
| Lupus nephritis | 37 |
| MN | 36 |
| TIN | 27 |
| MCNS | 26 |
| Diabetic nephropathy | 26 |
| Nephrosclerosis | 24 |
| Minor glomerular changes | 21 |
| FSGS | 19 |
| IgA vasculitis | 16 |
| TBM | 14 |
| MPGN | 11 |
| Amyloid Nephropathy | 8 |
| Alport syndrome | 5 |
| Obesity-related nephropathy | 4 |
| TMA | 4 |
| IRGN | 3 |
| Cryoglobulinemic vasculitis | 3 |
| Immunotactoid nephritis | 2 |
| Fabry disease | 2 |
| Non-IgA mesangial proliferative glomerulonephritis | 1 |
| Anti-GBM glomerulonephritis | 1 |
| Light-chain proximal tubulopathy | 1 |
| Medullary cystic kidney disease | 1 |
| Polyarteritis nodosa | 1 |
| Lipoprotein glomerulopathy | 1 |
| BK virus–associated nephropathy | 1 |

ANCA, anti-neutrophil cytoplasmic antibody; MN, membranous nephropathy; TIN, tubulointerstitial nephritis; MCNS, minimal change nephrotic syndrome; FSGS, focal segmental glomerulosclerosis; TBM, thin basement membrane disease; MPGN, membranoproliferative glomerulonephritis; TMA, thrombotic microangiopathy; IRGN, infection-related glomerulonephritis; GBM, glomerular basement membrane. TIN includes sarcoid nephropathy and IgG4-related kidney disease. TMA includes calcineurin inhibitor-associated nephropathy.

**Supplemental Table 3. Cox proportional hazards analysis of urine pH score.**

| | | Model 0<br>HR (95%CI) | Model 1<br>HR (95%CI) | Model 2<br>HR (95%CI) | Model 3<br>HR (95%CI) |
|---|---|---|---|---|---|
| Urine pH score | Score 0 | 1.28<br>(0.85-1.91) | 1.17<br>(0.78-1.76) | 1.36<br>(0.89-2.08) | 1.31<br>(0.86-2.01) |
| | Score 1 | Reference | Reference | Reference | Reference |
| | Score 2 | 0.60<br>(0.35-1.03) | 0.64<br>(0.37-1.11) | 0.61<br>(0.35-1.09) | 0.64<br>(0.36-1.14) |

Model 0: No adjustment factors. Model 1: Sex, Age (per 5 years). Model 2: Model 1 + Urinary protein, eGFR (per 10 mL/min/1.73m$^2$). Model 3: Model 2 + IFTA. HR, hazard ratio; 95%CI, 95% confidence interval; IFTA, interstitial fibrosis and tubular atrophy.

**Supplemental Table 4. Spearman's correlation coefficients among clinical and pathological variables**

| | Urine pH score | |
|---|---|---|
| | Correlation coefficient | p-value |
| Age | -0.13 | 0.003 |
| BMI | -0.12 | 0.010 |
| sBP | -0.11 | 0.014 |
| dBP | -0.04 | 0.41 |
| eGFR | 0.30 | <0.001 |
| HbA1c | -0.14 | 0.001 |
| U-P | 0.06 | 0.22 |
| IFTA | -0.25 | <0.001 |
| Medullary fibrosis | -0.14 | 0.001 |
| Medullary inflammatory cell infiltration | -0.09 | 0.048 |
| Medullary cast formation | -0.19 | <0.001 |

BMI, body mass index; sBP, systolic blood pressure; dBP, diastolic blood pressure; eGFR, estimated glomerular filtration rate; HbA1c, hemoglobin A1c; U-P, urinary protein excretion; IFTA, interstitial fibrosis and tubular atrophy.

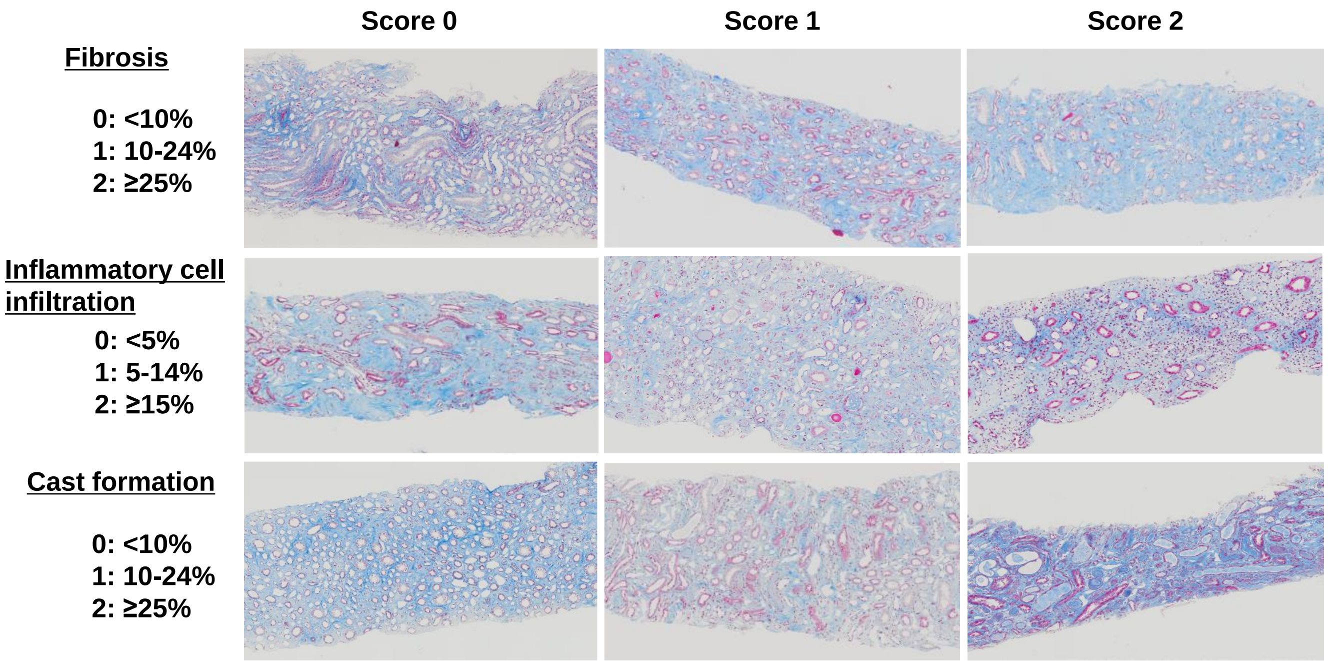


**Supplemental Figure 1. Medullary pathological findings and scoring system.**

Representative light microscopic images of renal medullary tissue stained with Masson's trichrome showing medullary fibrosis and cast formation. Each lesion was semi-quantitatively graded as 0, 1, or 2 based on severity.

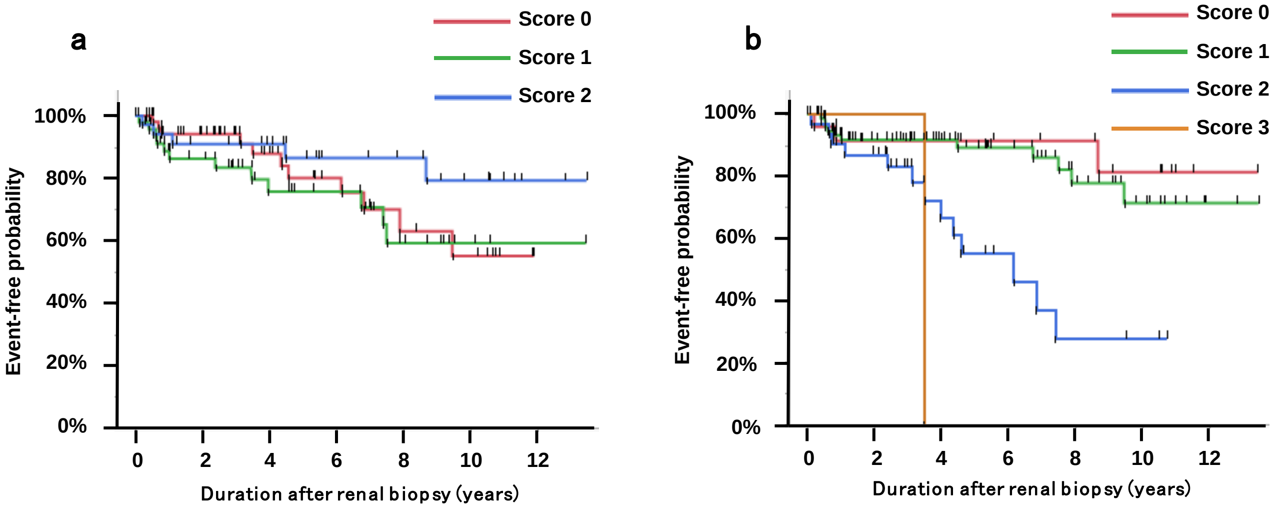


Number at risk

<Urine pH>

| | | | | | | |
|---|---|---|---|---|---|---|
| Score 0 | 68 | 40 | 26 | 17 | 9 | 7 |
| Score 1 | 50 | 33 | 19 | 15 | 9 | 3 |
| Score 2 | 40 | 27 | 23 | 15 | 13 | 9 |

Number at risk

<IFTA>

| | | | | | | |
|---|---|---|---|---|---|---|
| Score0 | 31 | 18 | 14 | 11 | 10 | 7 |
| Score1 | 90 | 56 | 42 | 30 | 18 | 10 |
| Score2 | 36 | 25 | 12 | 6 | 3 | 2 |
| Score3 | 1 | 1 | 0 | 0 | 0 | 0 |

**Supplemental Figure 2. Renal survival rate stratified by urine pH and IFTA score in IgA nephropathy.**

(a) Renal survival rate stratified by urine pH score. Log rank test: *p* = 0.241. (b) Renal survival rate stratified IFTA score. Log rank test: *p* <0.001. IFTA, interstitial fibrosis and tubular atrophy.

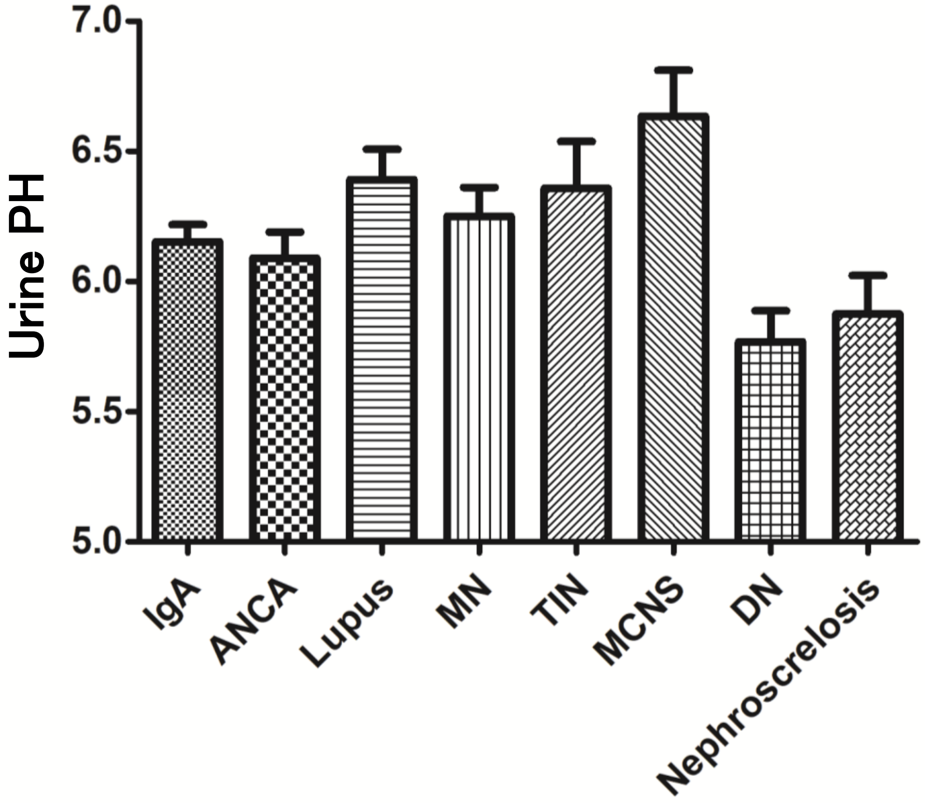


**Supplemental Figure 3. Urine pH according to underlying kidney disease.**

Urine pH according to underlying clinicopathological diagnoses in the study cohort. IgA nephropathy, ANCA-associated vasculitis, lupus nephritis, MN, TIN, MCNS, DN and nephrosclerosis. Boxes indicate median and interquartile range, with whiskers representing the range. ANCA, anti-neutrophil cytoplasmic antibody; MN, membranous nephropathy; TIN, tubulointerstitial nephritis; MCNS, minimal change nephrotic syndrome; DN, diabetic nephropathy.

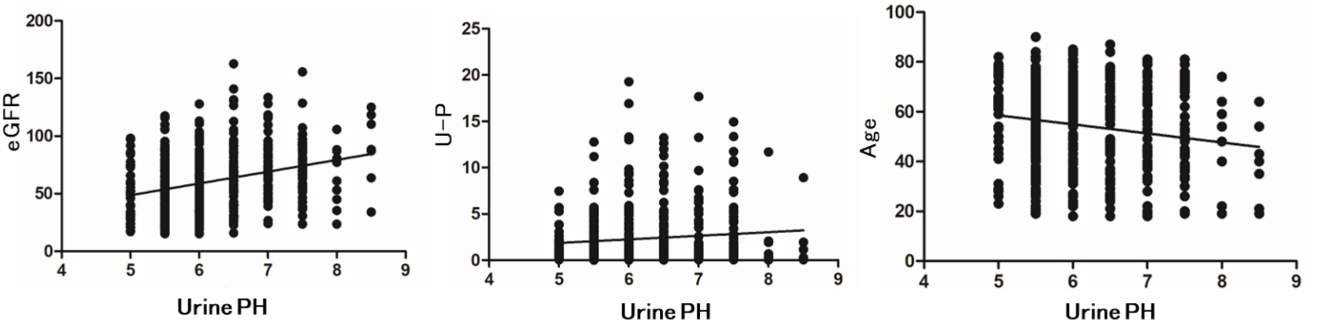


**Supplemental Figure 4. Correlation between urine pH and clinical variables.**
Scatter plots showing the relationships between urine pH and estimated glomerular filtration rate (eGFR), urine protein (U-P), and age at the time of kidney biopsy. Correlations were assessed using Spearman's rank correlation coefficient.

**STROBE Statement**

| | Item No | Recommendation | Page No |
|---|---|---|---|
| **Title and abstract** | 1 | (*a*) Indicate the study's design with a commonly used term in the title or the abstract | 1 |
| | | (*b*) Provide in the abstract an informative and balanced summary of what was done and what was found | 3-4 |
| **Introduction** | | | |
| Background/rationale | 2 | Explain the scientific background and rationale for the investigation being reported | 5-6 |
| Objectives | 3 | State specific objectives, including any prespecified hypotheses | 6 |
| **Methods** | | | |
| Study design | 4 | Present key elements of study design early in the paper | 7 |
| Setting | 5 | Describe the setting, locations, and relevant dates, including periods of recruitment, exposure, follow-up, and data collection | 7 |
| Participants | 6 | (*a*) Give the eligibility criteria, and the sources and methods of selection of participants. Describe methods of follow-up | 7 |
| | | (*b*) For matched studies, give matching criteria and number of exposed and unexposed | N/A |
| Variables | 7 | Clearly define all outcomes, exposures, predictors, potential confounders, and effect modifiers. Give diagnostic criteria, if applicable | 8-9 |
| Data sources/ measurement | 8* | For each variable of interest, give sources of data and details of methods of assessment (measurement). Describe comparability of assessment methods if there is more than one group | 8-9 |
| Bias | 9 | Describe any efforts to address potential sources of bias | 7-9 |
| Study size | 10 | Explain how the study size was arrived at | 7 |

| | | | |
|---|---|---|---|
| Quantitative variables | 11 | Explain how quantitative variables were handled in the analyses. If applicable, describe which groupings were chosen and why | 9-10 |
| Statistical methods | 12 | (*a*) Describe all statistical methods, including those used to control for confounding | 9-10 |
| | | (*b*) Describe any methods used to examine subgroups and interactions | 9-10 |
| | | (*c*) Explain how missing data were addressed | N/A |
| | | (*d*) If applicable, explain how loss to follow-up was addressed | N/A |
| | | (*e*) Describe any sensitivity analyses | N/A |
| **Results** | | | |
| Participants | 13* | (a) Report numbers of individuals at each stage of study—eg numbers potentially eligible, examined for eligibility, confirmed eligible, included in the study, completing follow-up, and analysed | 11 |
| | | (b) Give reasons for non-participation at each stage | Figure 1 |
| | | (c) Consider use of a flow diagram | Figure 1 |
| Descriptive data | 14* | (a) Give characteristics of study participants (eg demographic, clinical, social) and information on exposures and potential confounders | 11-12 |
| | | (b) Indicate number of participants with missing data for each variable of interest | |
| | | (c) Summarise follow-up time (eg, average and total amount) | |
| Outcome data | 15* | Report numbers of outcome events or summary measures over time | 11 |

| | | | |
|---|---|---|---|
| Main results | 16 | (*a*) Give unadjusted estimates and, if applicable, confounder-adjusted estimates and their precision (eg, 95% confidence interval). Make clear which confounders were adjusted for and why they were included<br>(*b*) Report category boundaries when continuous variables were categorized<br>(*c*) If relevant, consider translating estimates of relative risk into absolute risk for a meaningful time period | 11-14<br>9<br>N/A |
| Other analyses | 17 | Report other analyses done—eg analyses of subgroups and interactions, and sensitivity analyses | 12 |
| **Discussion** | | | |
| Key results | 18 | Summarise key results with reference to study objectives | 15 |
| Limitations | 19 | Discuss limitations of the study, taking into account sources of potential bias or imprecision. Discuss both direction and magnitude of any potential bias | 17 |
| Interpretation | 20 | Give a cautious overall interpretation of results considering objectives, limitations, multiplicity of analyses, results from similar studies, and other relevant evidence | 15-16 |
| Generalisability | 21 | Discuss the generalisability (external validity) of the study results | 17 |
| **Other information** | | | |
| Funding | 22 | Give the source of funding and the role of the funders for the present study and, if applicable, for the original study on which the present article is based | 18 |
